# WEIRD or not: A Cross-Cultural Behavioral Economic Assessment of Demand for HIV and COVID-19 Vaccines

**DOI:** 10.1101/2023.07.24.23293101

**Authors:** Promise Tewogbola, Eric A. Jacobs, Justin T. McDaniel

**Affiliations:** School of Psychological and Behavioral Sciences Southern Illinois University, Carbondale, IL; School of Human Sciences, Southern Illinois University, Carbondale, IL

**Keywords:** HIV vaccines, COVID-19 vaccines, behavioral economics, behavioral economic demand, cross-cultural

## Abstract

**Background:** Despite empirical evidence supporting vaccine effectiveness, vaccine hesitancy continues to thrive. Demand as a behavioral economic process provides useful indices for evaluating vaccine acceptance likelihood in individuals and groups. Using this framework, our study investigates the dynamics governing vaccine acceptance in two culturally dissimilar countries.

**Methods:** Hypothetical purchase tasks (HPTs) assessed how Nigerian and US participants varied vaccine acceptance as a function of hospitalization risks due to vaccination (*N* = 109). Aggregate and individual demand indices (*Q*_0_ and *P*_max_) were computed with nonlinear regressions. Secondary analyses were conducted using repeated measures ANOVAs with vaccine type (COVID-19 and HIV) as the within-subject factor; country, age, and socioeconomic status as between-subjects factors; demand indices served as dependent variables.

**Results:** Demand indices varied significantly as a function of vaccine type (*F* (1, 57) = 17.609, *p <* .001, 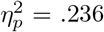). Demand for HIV vaccines was higher relative to COVID19 vaccines. Interactions between vaccine type and country of origin (*F* (1, 56) = 4.001, *p* = .05, 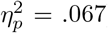) were also significant with demand for HIV vaccines among Nigerian respondents higher than that of COVID-19 vaccines. This was reversed for US participants. Interactions between vaccine type, country of origin and age were also significant (*F* (2, 51) = 3.506, *p <* .05, 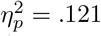).

**Conclusions:** These findings provide evidence that vaccine type can influence demand. The relationship between demand and vaccine type also varies as a function of country of origin and age. Significance, limitations, and future directions are also discussed.

## 1 Introduction

Vaccines are a remarkable innovation in the healthcare industry, responsible for saving millions of lives each year (Habersaat & Jackson, 2020). These medical marvels offer immunity from a wide range of illnesses, including COVID-19, tetanus, measles, rabies, smallpox, diphtheria, and others (Ellenberg & Chen, 1997; Prüß, 2021). Despite the well-known benefits of vaccines, people show reluctance or delay in accepting them. Vaccine hesitancy continues to be a widespread issue globally and can be driven by various factors, including a lack of knowledge, religious beliefs, political affiliations, and complacency (Yaqub et al., 2014; Pelcic et al., 2016; Karafillakis & Larson al., 2017; Olagoke et al., 2021; Fridman et al., 2021). As noted by d’Onofrio and Mafredi (2010), vaccine hesitancy can also stem from a perception of increased hospitalization risks due to accepting a vaccine. In this context, vaccine hesitancy can be assessed as it relates to the efficacy of the vaccines in preventing hospitalization. By reframing this in terms of behavioral economic demand, the dynamics governing vaccine-related behavior, whether acceptance or hesitancy, can be better understood.

Behavioral economics is a field that leverages economic principles and psychological insights to comprehend the people’s decision-making process (Angner & Loewenstein, 2012; Hursh & Roma, 2013; Thaler, 2018). Within this framework, demand is the relationship between the consumption of a commodity and its unit price, which can be represented by a non-linear demand curve (Allen, 1962; Jacobs & Bickel, 1999). Figure 1 shows a schematic representation of such demand curve. The price of choosing a health-related commodity, however, is not limited to its monetary cost. The time and effort involved in getting to a vaccine, the social costs of stigma associated with vaccine acceptance, and the effort of remembering to attend one’s vaccination appointment are all examples of costs that are not necessarily financial in nature. Uncertainty and risk are also other forms of cost within the behavioral economic framework because the decision-making agent sometimes need to make choices in the absence of clear-cut discriminating stimuli in the environment (Altman, 2012). In all, an analysis of the demand curves of different health commodities can reveal information about how these commodities are valued by the consumer. This is called the willingness-to-pay (WTP) approach to demand analysis (Phelps, 2010).

**Figure 1:**
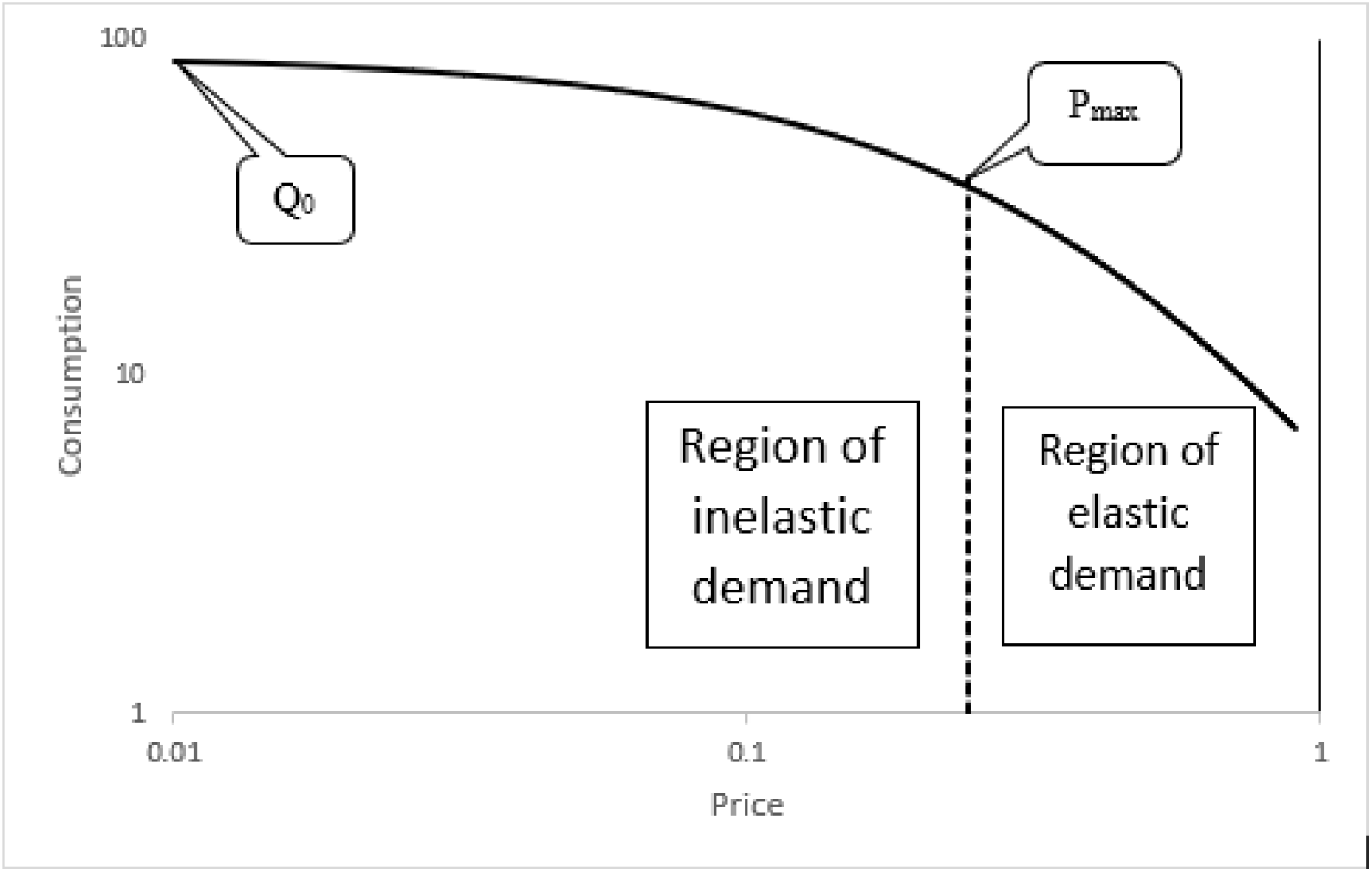
Schematic exponential demand curve plotted on logarithmic axes. *Q*_0_ depicts the highest level of consumption when the commodity is free. Demand transitions from inelastic to elastic at *P*_max_ which is the point where the slope is −1

The hypothetical purchase task (HPTs) is a widely used method to assess demand and derive demand curves for commodities such as alcohol, marijuana, heroin, gym memberships, condoms, and cigarettes (Gentile et al., 2012; Collins et al., 2014; Jacobs & Bickel, 1999; MacKillop et al., 2008; Brown et al., 2021; Strickland et al., 2020). Support for the use of the HPTs as a behavioral economic tool is provided by the evidence of correspondence between performance for simulated outcomes in the HPTs and real outcomes (see Amlung et al., 2012). This is in addition to the reduced financial, temporal and effort costs associated with the use of the HPT as opposed to using real outcomes in demand analysis experiments.

Demand can be mathematically modelled with an exponential function (Hursh & Silberberg, 2008) which is given as:

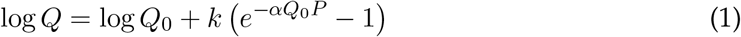

where *Q* represents the quantity of the commodity of interest consumed, and *Q*_0_ depicts the highest level of demand when that commodity is free. The parameter, *α*, specifies the rate of decline in relative consumption with increases in cost. *P* stands for price, and *k* is the scaling constant reflecting the range of consumption data in log units.

Also relevant to the discussion on behavioral economic demand is the concept of elasticity, which refers to the changes in consumption which are a function of the changes in cost-benefit ratio (Gilroy et al, 2019). In other words, it reflects the sensitivity of demand for a commodity to changes in price, with some regions of the demand curve showing high sensitivity to price changes and others showing relative insensitivity. Behavioral economic studies of people who use illicit drugs, for instance, have shown that highly valued drugs are insensitive and inflexible to changes in price. This can result in addicted individuals incurring disproportionate costs (in terms of time, money, or effort) for these harmful commodities (Petry & Bickel, 1998; Payne et al., 2020).

Elasticity of demand can be categorized as elastic, inelastic, or unit elastic, depending on the relationship between price changes and subsequent changes in consumption. Inelastic demand means that consumption levels remain constant despite large changes in price, whereas elastic demand means that slight changes in cost result in proportionally larger changes in consumption (Madden, 2000). The point of unit elasticity, which corresponds with the maximum price (*P*_max_), indicates the transition from inelastic to elastic demand on the demand curve (see Figure 1).

The highest level of demand (*Q*_0_) and the point of transition from inelastic to elastic demand (*P*_max_) are two of the most important indices used in comparing demand between individuals or across population groups. In the context of vaccines, higher *Q*_0_ and *P*_max_ values are both indicative of a greater subjective valuation of the vaccine, albeit in different ways. A high *Q*_0_ indicates a strong likelihood of accepting a vaccine when the cost is low or zero, while a high *P*_max_ suggests that the respondent is willing to tolerate a high cost in order to receive a vaccine.

Consequently, the aim of our study is three-fold. First, we will assess how vaccine demand varies as a function of vaccine type. The types of vaccines that will be considered are of two varieties the more recently developed and widely available COVID-19 vaccine and the HIV vaccine which is still in development for mass utilization. Assessing vaccine demand, particularly that of HIV vaccines, within a behavioral economic framework is expected to have public health significance – especially for population groups most at risk of getting infected with HIV.

Second, we will explore how vaccine demand varies as a function of geographical location. Participants were recruited from the US, which is the prototypical Western, educated, industrialized, rich, and democratic (WEIRD) nation and Nigeria, an exemplar of the non-WEIRD country often not the subject of behavioral economic studies (Henrich et al., 2010). We will be investigating how the dynamics of vaccine demand in the two countries are different.

Finally, we will be examining the interactions between vaccine type, geographical location, and other personal characteristics such as age and socioeconomic status.

## 2 Methodology

### 2.1 Sampling Procedure

Our study recruited participants from both the United States and Nigeria through online platforms. American participants were obtained through Amazon’s crowdsourcing platform, Mechanical Turk (mTurk), while Nigerian participants were sourced from social media websites. Participants from both countries were informed and provided with informed consent before taking part in the study questionnaire. A final sample of 109 respondents was analyzed after removing participants who did not provide systematic data or showed an increasing likelihood of accepting vaccines at higher risks of hospitalization. The study was approved by the Southern Illinois University Institutional Review Board (IRB) and all procedures were in accordance with ethical guidelines. American participants were compensated with $1, while Nigerian participants were uncompensated.

### 2.2 Vaccine Demand Procedure

Vaccine demand was evaluated through two different hypothetical purchase tasks – one for COVID-19 and the other for HIV. Participants were presented with vignettes adapted from Hursh and colleagues’ (2020) study. The vignettes indicated that the vaccines were available for free, would have to be administered immediately, and had been approved by either the Food and Drug Administration (FDA) for US participants or the National Agency for Food and Drug Administration and Control (NAFDAC) for Nigerian participants. All respondents were asked to report the likelihood of accepting COVID-19 and HIV vaccines based on different levels of vaccine efficacy, which we defined as the degree to which the vaccine reduces the chances of disease-related hospitalization. This was presented on a scale ranging from 100% (the vaccine is completely effective at preventing disease-related hospitalization) to 0% (the vaccine does not prevent disease-related hospitalization) in decrements of 10 percentage points. Participants’ willingness to accept a vaccine was then measured on a different scale from 0% (completely unwilling to be vaccinated) to 100% (completely willing to be vaccinated).

### 2.3 Demographics

Upon completing the COVID-19 and HIV vaccine purchase tasks, participants also provided other demographic information such as age, gender, subjective socioeconomic status and country of origin.

### 2.4 Data Analyses

A nonlinear regression was used to compute individual demand curves for each participant, as well as the aggregate demand curve for all participants. The exponential demand function (Hursh & Silberberg, 2008) was fit to the demand curve representing the relationship between each participants’ likelihood of accepting a vaccine (*Q*) and the varying risks of future hospitalization (*P*). Likelihood values for all participants were expressed in percentages (0 - 100%) while risks of future hospitalization were converted to probabilities ranging from 0 (no risk of future hospitalization) to 1 (certain hospitalization in the future). Values for *Q*_0_ (likelihood of vaccine acceptance at zero risk) and *α* (rate in decline of likelihood to accept vaccine) were parameters generated by the exponential demand model – although, *Q*_0_ values were also empirically collected in the study. The model-generated *Q*_0_ values were constrained to 100% as the likelihood of vaccine acceptance ranged from 0% to 100%. Furthermore, since the logarithm of zero is undefined, when empirical vaccine acceptance likelihood values were 0%, they were replaced with 0.001%. The goodness of fit for each participant’s demand curve was assessed through the percentage of variance accounted for (*R*^2^). Table 1 shows the *Q*_0_ and *α* values for the top 10 best fitting individual curves in the COVID-19 and HIV conditions. The Lambert W function (Gilroy et al., 2019) was used to calculate the *P*_max_ by utilizing the empirical *Q*_0_ and the model-generated *α*:

**Table 1:** Parameter Estimates for COVID-19 and HIV Vaccines with the Top 10 Best Model Fits.

| Vaccine | $Q_0$<br>(%) | $\alpha$ | $R^2$ |
| --- | --- | --- | --- |
| COVID-19 | 10.298 | -.041 | 0.974 |
|  | 66.306 | 0.007 | 0.966 |
|  | 66.783 | 0.009 | 0.956 |
|  | 100.000 | 0.003 | 0.940 |
|  | 58.630 | 0.007 | 0.940 |
|  | 60.866 | 0.004 | 0.934 |
|  | 71.224 | 0.002 | 0.918 |
|  | 7.568 | -.065 | 0.910 |
|  | 100.000 | 0.004 | 0.899 |
|  | 100.000 | 0.006 | 0.896 |
| HIV | 61.459 | 0.003 | 0.961 |
|  | 85.268 | 0.001 | 0.958 |
|  | 70.030 | 0.006 | 0.957 |
|  | 95.087 | 0.008 | 0.955 |
|  | 71.405 | 0.008 | 0.948 |
|  | 23.532 | -.013 | 0.939 |
|  | 100.000 | 0.001 | 0.939 |
|  | 56.595 | 0.007 | 0.920 |
|  | 61.660 | -.001 | 0.915 |
|  | 100.000 | 0.002 | 0.911 |

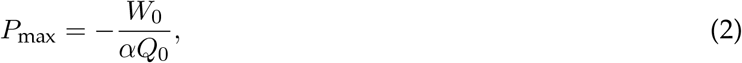

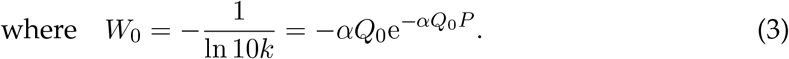

Negative *P*_max_ values were recorded as 0.001, while those that were equal to or greater than 1 were recoded as 0.99 since the risk of future hospitalization on account of vaccine ineffectiveness can neither be negative nor greater than 1.

The *P*_max_ and empirical *Q*_0_ values both served as the dependent variables. Empirical, rather than the model-derived *Q*_0_, values were used as the dependent variable because all participants were directly asked about their likelihood of accepting a vaccine at zero risk of hospitalization. However, the strong correlations between the empirical and modelderived *Q*_0_ values lend credence to the validity of the former as a suitable dependent variable (see Table 2).

**Table 2:** Correlations between Empirical (E) and Model-derived (MD) *Q*_0_. All correlations were statistically significant at *p <* 0.001.

| | E-$Q_0$ COVID-19 | E-$Q_0$ HIV | MD-$Q_0$ COVID-19 | MD-$Q_0$ HIV |
| --- | --- | --- | --- | --- |
| E-$Q_0$ COVID-19 | — | | | |
| E-$Q_0$ HIV | 0.787 | — | | |
| MD-$Q_0$ COVID-19 | 0.846 | 0.673 | — | |
| MD-$Q_0$ HIV | 0.564 | 0.668 | 0.622 | — |

A series of analyses of variance (ANOVAs) were used to evaluate how *P*_max_ and the empirical *Q*_0_ varied as a function of (1) vaccine type (COVID-19 or HIV); (2) country of origin (Nigeria or USA); and (3) interactions between vaccine type, country of origin, and other personal characteristics. All analyses were conducted with SPSS v. 23.

## 3 Results

### 3.1 Demographics

Among the 109 respondents, about 58% of the participants were female, while almost 40% were Americans. A little less than half (46.8%) were aged between 25 and 34, while more than 50% of the participants categorized themselves as middle-to-upper income earners. Demographic data is summarized in Table 3.

**Table 3:** Participants’ Demographic Information.

| Category | Subcategory | Frequency | Percentage |
| --- | --- | --- | --- |
| Gender | Male | 44 | 40.4 |
|  | Female | 63 | 57.8 |
| Age | 18–24 | 30 | 27.5 |
|  | 25–34 | 51 | 46.8 |
|  | 35+ | 24 | 22.0 |
| Country | Nigeria | 65 | 59.6 |
|  | USA | 44 | 40.4 |
| Socioeconomic status | Lower | 49 | 45.0 |
|  | Middle-to-Upper | 58 | 53.2 |

### 3.2 Aggregate Demand for Vaccine

At the aggregate level, the likelihood of vaccine acceptance generally decreased as the risk of hospitalization on account of vaccination increased. The exponential demand model provided a good fit for the aggregate demand values for COVID-19 (*R*^2^ = 0.92) and HIV (*R*^2^ = 0.87) vaccines. The aggregate model-derived *Q*_0_ values for COVID-19 and HIV vaccines were 91.2% and 92.4%, respectively. On the other hand, the aggregate *P*_max_ value for COVID-19 vaccines was 0.54, while that of HIV vaccines was 0.70. The aggregate demand curves for COVID-19 and HIV vaccines are shown in Figure 2.

**Figure 2:**
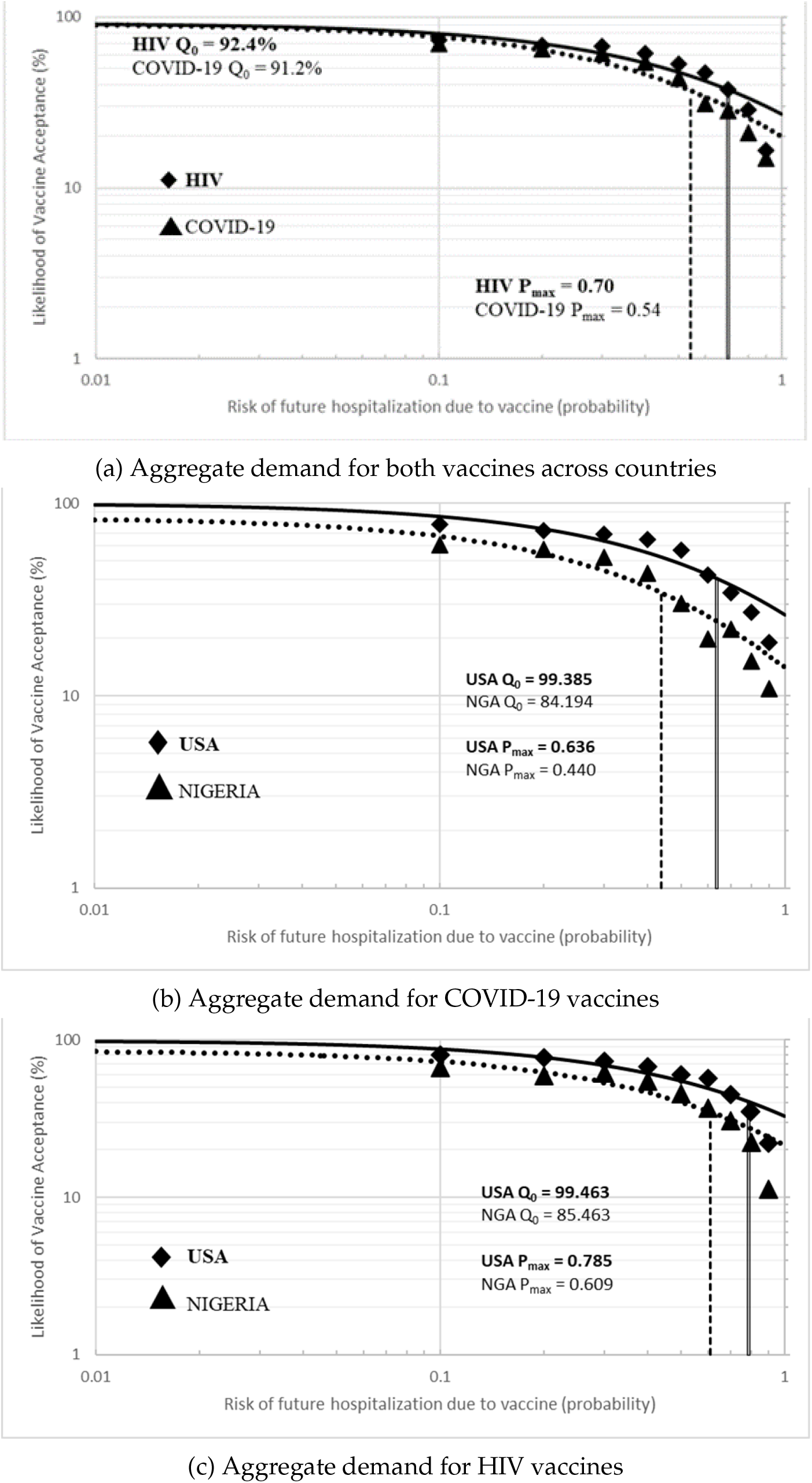
Aggregate Demand Curves for COVID-19 and HIV vaccines plotted on logarithmic scales. *Note:* In Figure 2a, the indices of aggregate demand for all countries is higher for HIV vaccines compared to COVID-19 vaccines; Figure 2b and 2c, show that aggregate demand for both vaccines is higher in the US compared to Nigeria

### 3.3 Correlations between Empirical and Model-derived *Q*_0_

In order to evaluate the strength of the relationship between the empirical and modelderived *Q*_0_ values, a Pearson’s correlation coefficient was computed. For COVID-19 vaccines, empirical and model-derived *Q*_0_ values were found to be strongly positively correlated, *r*(107) = .846, *p <* .001. A strong positive correlation was also obtained for HIV vaccines, *r*(107) = .673, *p <* .001. Table 2 shows the correlations between the empirical and model-generated *Q*_0_ values.

### 3.4 Vaccine Demand by Vaccine Type

A within-subjects repeated measures ANOVA was conducted to investigate the effects of vaccine type (COVID-19 or HIV) on empirical *Q*_0_ and *P*_max_ values. There was a significant difference between the empirical *Q*_0_ for COVID-19 and HIV (*F* (1, 57) = 17.609, *p <* .001, 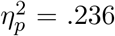). The average empirical *Q*_0_ for COVID-19 was 79.9%, which increased to 87.4% for HIV – a difference of 7.5 percentage points (*SD* = 1.788). On the other hand, the average *P*_max_ values for HIV was 0.572 while that of COVID-19 was 0.508 – a difference of 0.064. However, the main effect of vaccine type on *P*_max_ values was marginally nonsignificant (*F* (1, 57) = 3.903, *p* = .053, 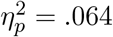).

### 3.5 Vaccine Demand by Country

Empirical *Q*_0_ and *P*_max_ values were analyzed using separate 2 × 2 mixed ANOVAs with country as the between-subject factor (Nigeria or USA) and vaccine type as the within-subject factor (COVID-19 and HIV). The average empirical *Q*_0_ values for Nigerian participants were not statistically different from that of American participants (*F* (1, 56) = 0.867, *p* = .356, 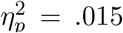). The *P*_max_ values acceptable to American respondents were 0.617, while those of Nigerian respondents were 0.49 – a difference of 0.127. The contrast on this difference, however, was not statistically significant (*F* (1, 56) = 3.81, *p* = .056, 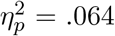). That said, the interaction between vaccine type and country was significant for *P*_max_ values (*F* (1, 56) = 4.001, *p* = .05, 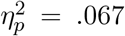). For Nigerian participants, the *P*_max_ values for COVID-19 vaccines (*M* = 0.433, *SD* = 0.041) were lower than those of HIV vaccines (*M* = 0.547, *SD* = 0.05). This relationship was reversed for American participants, with the *P*_max_ values for COVID-19 vaccines higher (*M* = 0.624, *SD* = 0.051) than those of HIV vaccines (*M* = 0.61, *SD* = 0.062). The graphs comparing the means of *Q*_0_ and *P*_max_ values for Nigerian and American participants are shown in Figure 3.

**Figure 3:**
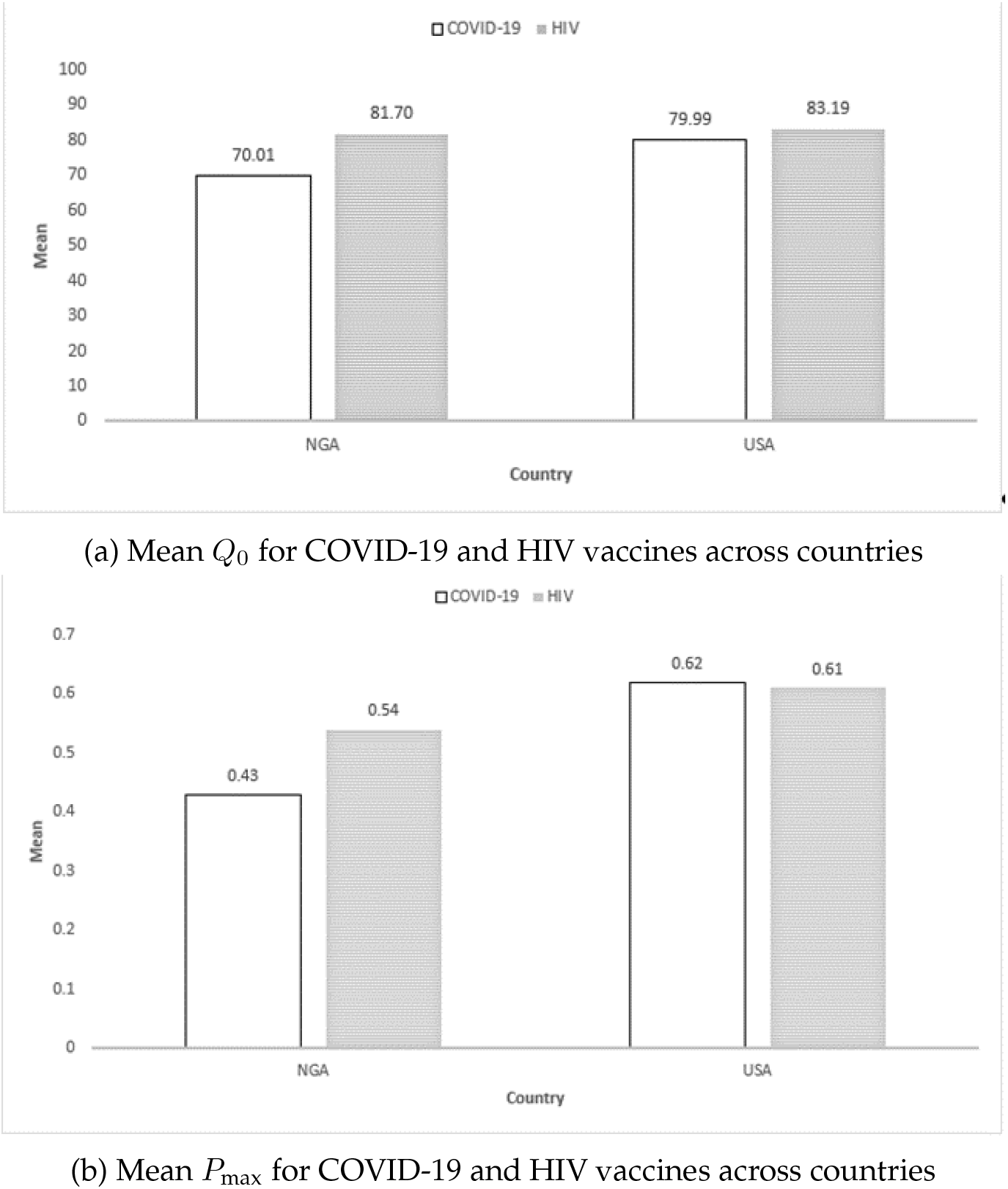
Differences in mean empirical *Q*_0_ and *P*_max_ values for COVID-19 and HIV vaccines as a function of country of origin. *Note:* In Figure 3a, *Q*_0_ for both HIV and COVID-19 vaccines are higher for the US compared to Nigeria; Figure 3b, on the other hand, *P*_max_ values show that aggregate demand for HIV vaccines are higher than COVID-19 vaccines in Nigeria, while the reverse is the case in the US

### 3.6 Vaccine Demand by Interactions between Country and Other Personal Characteristics

Secondary analyses of empirical *Q*_0_ and *P*_max_ values were conducted through 2 separate three-way mixed ANOVAs. Aside from country of origin and vaccine type, the other additional factors considered for analysis were age (“18 – 24”, “25-34” and “35+”) and socioeconomic status (“Lower” and “Middle-to-Upper”).

#### 3.6.1 Country by Age by Vaccine Type

Although there were no statistically significant differences between the empirical *Q*_0_ of respondents in different age groups (*F* (2, 51) = 1.478, *p* = .238, 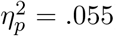), the interaction between country, age, and vaccine type was significant (*F* (2, 51) = 3.506, *p <* .05, 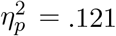). More specifically, for Nigerian participants aged 24 – 34, the empirical *Q*_0_ for HIV vaccines was lower (*M* = 81.76, *SD* = 5.32) than that of their counterparts from the USA (*M* = 83.48, *SD* = 6.63). This situation was reversed among participants aged 35 and above, with the empirical *Q*_0_ for HIV vaccines higher for Nigerians (*M* = 98.89, *SD* = 11.47) in comparison to Americans (*M* = 91.02, *SD* = 5.52). Similarly, in the 24 – 34 group, Americans’ empirical *Q*_0_ values for COVID-19 vaccines were much higher (*M* = 80.83, *SD* = 7.143) than that of Nigerian participants (*M* = 68.95, *SD* = 5.73) – a difference of 14.53 percentage points. In the 35+ group, however, the difference between American (*M* = 89.64, *SD* = 5.94) and Nigerian (*M* = 87.22, *SD* = 12.37) participants reduced to 2.42 percentage points. On the other hand, the main effect of age on *P*_max_ values was not statistically significant (*F* (2, 51) = 2.047, *p* = .140, 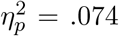). Interactions were also not significant (*F* (2, 51) = 0.184, *p* = .833, 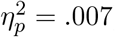).

#### 3.6.2 Country by Socioeconomic Status by Vaccine Type

Neither the main effects of socioeconomic status (*F* (1, 53) = 2.554, *p* = .116, 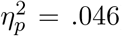), nor its interactions with country and vaccine type were statistically significant for the empirical *Q*_0_ (*F* (1, 53) = 0.014, *p* = .906, 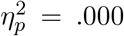). Similarly, socioeconomic status had no significant effect on *P*_max_ values (*F* (1, 53) = 0.657, *p* = .421, 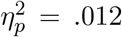). Its interactions with country and vaccine type were also nonsignificant (*F* (1, 53) = 1.892, *p* = .175, 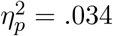).

## 4 Discussion

The present study explores the dynamics driving demand for vaccines through the lens of behavioral economics. This examination is novel in its approach, as it is the first to assess the demand for HIV vaccines in this manner. The ramifications of an effective HIV vaccine cannot be underestimated. A preventive HIV vaccine offers the prospect of lifelong protection and tackles the prevalent challenges associated with HIV treatment, including adverse side effects, inadequate adherence, and stigma (Duncan et al., 2012; Johnson & Neilands, 2007; Van Tam et al., 2011). Moreover, a successful HIV vaccine presents a more cost-effective alternative to a lifetime of antiretroviral therapy and prophylactics.

In March 2022, the National Institute of Allergy and Infectious Diseases (NIAID) initiated clinical trials evaluating three experimental HIV vaccines based on the same mRNA technology used in various approved COVID-19 vaccines (Harris, 2022; Rogers, 2022). These trials – which, at present, have enrolled 108 HIV-uninfected adults (U.S. National Library of Medicine, 2022) –, with Dr. Anthony Fauci, the Director of NIAID, commenting, “HIV research absolutely helped COVID-19. Now that we have a successful vaccine with mRNA, it’s going to go back. Everything that goes around comes around. We’re going to hopefully get more insight into HIV vaccines.” (Stulpin, 2021). The advent of a viable HIV vaccine is imminent and has the potential to save millions of lives across the globe.

The study at hand represents a pioneering effort in utilizing behavioral economic analysis to shed light on the demand for vaccines in Nigeria. Although the tools and methods of behavioral economics can provide an understanding of choice and decision making in a variety of domains, empirical grounding of behavioral economic recommendations can be strengthened through the examination of diverse subject pools. Despite the abundance of cross-cultural studies in behavioral economics (e.g., Chen, 2013; Doces & Wolaver, 2021; Domino, 1992; Henrich et al., 2010; Levinson & Peng, 2007; Wright & Phillips, 1980; Yates, et al., 2002), few have focused on the domain of health behavior. The present study bridged this knowledge gap by highlighting how the demand for HIV and COVID-19 vaccines vary as a function of cultural differences.

Regardless of the vaccine type, the findings of this study showed that vaccine demand generally decreased as the risk of hospitalization on account of vaccination increased. That said, when vaccines were deemed to be 100% effective (*Q*_0_), there was a significant difference between the vaccine acceptance likelihood for COVID-19 vaccines and that of HIV vaccines, with the demand for HIV vaccines higher than that of COVID-19 vaccines. However, on aggregate, the maximum acceptable risk of future hospitalization (*P*_max_) was similar for both HIV and COVID (ranging between 50 – 57%). Thus, at the aggregate level of analysis, the type of vaccine appears to influence its demand, but not its maximum acceptable perceived risk of future hospitalization from that vaccine.

Our finding that demand for COVID-19 vaccines was lower than that of HIV vaccines was unexpected. This runs counter to the predictions of cognitive biases such as recency bias and the availability heuristic (Tversky & Kahneman, 1973), which suggest that demand for the COVID-19 vaccine should be higher due to its recent prominence in the public consciousness, as opposed to HIV which was first clinically reported in the 1980s (Gottlieb et al., 1981; Wu et al., 2020). These results imply that some other factor is driving the higher demand for HIV vaccines. One possible explanation is the high recovery rate of COVID-19 cases. Unlike HIV and other debilitating viral diseases, there are numerous cases of people getting infected with COVID-19 and later recovering (Tewogbola & Aung, 2021). According to the Worldometer Coronavirus Dashboard (2022), up to 99% of infected individuals recover. Although some individuals may suffer from long-term symptoms like fatigue, dyspnea, and cognitive impairment, known as “long COVID,” the vast majority of COVID-19 cases are resolved (Crook et al., 2021). About a third of those infected with COVID-19 remain asymptomatic while severe illnesses occur only in a small proportion of the symptomatic cases (Doshi, 2020; Gao et al., 2021; Oran & Topol, 2021). On the other hand, only a small percentage of individuals infected with HIV have a clinically undetectable viral load without undergoing antiretroviral therapy (ART) (Blankson, 2010; Grabar et al., 2009). Unlike COVID-19, the median survival time for an individual infected with HIV who does not undergo treatment is 9.2 months (Morgan et al., 2002). Given these considerations, it is possible that individuals may be less motivated to seek protection from a disease that is perceived to have a high rate of recovery and a low likelihood of severe illness. In contrast, HIV is widely recognized as a debilitating and life-threatening disease, which may drive higher demand for its vaccine.

Our study sheds new light on the differing attitudes towards risk acceptance for vaccines between Nigeria and the United States. Our findings reveal that Nigerian participants were willing to accept a higher maximum risk of future hospitalization for an HIV vaccine than for a COVID-19 vaccine, whereas American participants showed the opposite preference. This result aligns with the current epidemiology of the two diseases in each country. According to the WHO estimates (2022), Nigeria experiences approximately 74,000 new cases of HIV each year, compared to 34,800 in the United States. On the other hand, the weekly incidence rate of COVID-19 in the US was almost 280,000 cases in February 2023, compared to the complete absence of reported new cases in Nigeria (WHO, 2023). These disparities in disease prevalence appear to inform the differing levels of risk tolerance among participants in each country. It is clear that the risk of future hospitalization associated with a vaccine is perceived differently based on the local incidence of the disease being targeted.

The results of our study also suggest that religiosity may play a role in COVID-19 vaccine acceptance. Previous research has indicated a correlation between religiosity and vaccine acceptance, with more religious individuals being less likely to accept a vaccine (Olagoke et al., 2021; Sallam, 2021). Given that African countries tend to have higher levels of religiosity compared to the United States (Stavrova et al., 2013), it is possible that this difference in religiosity may be reflected in demand for the COVID-19 vaccine. Similarly, a Pew Research report (2021) found that only 29% of Americans identify as irreligious, whereas only 0.4% of Nigerians report not being religious (McKinnon, 2021). These findings suggest that the lower degree of religiosity in the United States may contribute to the higher relative demand for COVID-19 vaccines. However, it is important to note our study did not directly measure religiosity and, therefore, further investigation is required to solidify this interpretation of the data.

Like the findings of Hursh and his collaborators (2020), we did not find a significant effect of socioeconomic status on vaccine demand. However, we did observe intriguing interactions between vaccine type, country of residence, and age. For participants in the 24 - 34 age group, the demand for HIV vaccines was lower among Nigerian participants compared to American participants. However, for those in the 35+ age group, the demand for HIV vaccines was higher among Nigerians compared to Americans. This could be due to the fact that the younger individuals in the 35+ age group were likely old enough to have experienced the massive HIV prevention programs that took place in Nigeria in the early 2000s (UNAID, 2006). These programs may have played a role in the higher demand for HIV vaccines relative to COVID-19 vaccines among older Nigerian participants in our study. While our findings offer a preliminary understanding of how factors interact to influence vaccine demand, further research is needed to fully explore the potential presence of other confounding variables.

Overall, the findings of this study present a comprehensive examination of the anticipated demand for crucial vaccines in both the United States and Nigeria. As research on the risk of hospitalization from vaccines, such as the HIV vaccine, continues to unfold, these results can serve as a valuable resource for healthcare practitioners in their planning and allocation of resources. The data provided in this study can assist public health professionals in Nigeria in determining the necessary amount of HIV vaccines required to cover the population. The challenge of efficiently allocating scarce healthcare resources (Emanuel et al., 2020) is a pressing issue, and having access to accurate demand data is crucial in overcoming this challenge. This study provides a valuable contribution to the field by illuminating the demand for critical vaccines, which can inform future resource allocation decisions.

Our study should be interpreted in light of its strengths and limitations. One strength is its use of the hypothetical purchase tasks (HPTs) to safely model demand for a novel and as-yet-unexperienced health commodity such as an HIV vaccine. In addition, HPTs, unlike the discrete dichotomous approach (e.g., Question: “Would you accept an HIV vaccine?”; Answer: “Yes” or “No”), are more robust to overestimation and underestimation of demand because they offer experimenters the ability to capture, isolate, and control for other factors facilitating the differences in demand between and within subjects (Strickland et al., 2022). However, our study also has some limitations that must be considered when interpreting the results. One such limitation is the use of non-probability sampling, as American participants were recruited from a crowdsourced platform, while Nigerian participants were recruited from social media sites. As noted by Chandler and Shapiro (2016), non-probability sampling can introduce selection bias and threaten the validity of the results. Another limitation was the different levels of familiarity participants had with the COVID-19 and HIV vaccines. Given that COVID-19 vaccines are globally distributed and extensively covered in the media, participants likely had direct experience or at least more familiarity with them, influencing their perceived demand in our hypothetical purchase task. In contrast, their experiences with the HIV vaccine were entirely hypothetical, which could have influenced their responses in ways we were not able to control. This interpretive difficulty complicates the direct comparison between demand for the two vaccine types. Therefore, results should be interpreted with this consideration in mind. Additionally, the relatively small sample size also limits the generalizability of our findings. Despite these limitations, the HPT method used in our study provides a robust assessment of demand for vaccines, offering valuable information for public health professionals and policymakers as they work to allocate scarce health resources and plan for the purchase and distribution of vaccines.

## 5 Conclusion

Taken together, the present study sheds light on the sensitivity of vaccine demand, as expressed by the likelihood of accepting a vaccine, to vaccine type and country of origin. Our results indicate that demand for HIV vaccines was higher overall than that for COVID-19 vaccines, suggesting that there might be less hesitancy towards accepting HIV vaccines when they are eventually made available. American participants showed a greater demand for COVID-19 vaccines compared to HIV vaccines, whereas Nigerian participants demonstrated the opposite pattern. It is worth noting that this study represents a pioneering effort in conducting a behavioral economic analysis of vaccine demand for HIV vaccines, and in using a Nigerian sample to investigate health decision-making from a behavioral economic perspective. Future research should focus on the impact of real and perceived risks of hospitalization on vaccine demand, as well as exploring demand for vaccines among populations most susceptible to certain diseases. For instance, conducting a behavioral economic analysis of vaccine acceptance likelihood among minority groups in the US or among men who have sex with men (MSMs) could provide valuable insights with significant public health and policy implications. Additionally, investigating how vaccine demand is influenced by other cost dimensions, such as the monetary price of vaccines or waiting time for vaccination appointments, is can also a promising direction for future research.

## Data Availability

We confirm that all data referenced in this manuscript are available upon reasonable request. Interested researchers may contact the corresponding author to access the datasets used and analyzed during the course of this study. We are committed to promoting transparency and reproducibility in our research, and we welcome inquiries to facilitate further exploration and verification of our findings.

## Notes

### Competing Interest Statement

The authors have declared no competing interest.

### Funding Statement

This study did not receive any funding.

### Author Declarations

The Institutional Review Board (IRB) of Southern Illinois University gave ethical approval for this work.

